# Universal Periodic Review recommendations and trajectories of maternal health between 2005 and 2023: a longitudinal ecological analysis of 89 countries

**DOI:** 10.64898/2026.06.03.26354800

**Authors:** Anshu Uppal, Rebekah Thomas, Mattia De Pasquale, Jesse Sillo, Haileyesus Getahun

## Abstract

**Background:** The Universal Periodic Review (UPR) is a peer-review mechanism established to hold UN Member States accountable for human rights including the right to health, yet evidence on its impact on health outcomes is limited. We evaluated whether UPR engagement is associated with accelerated improvements in maternal health trajectories.

**Methods and Findings:** We conducted a longitudinal ecological analysis of 89 countries with a baseline maternal mortality ratio (MMR) of 70 or greater per 100,000 live births in 2005. Outcomes were trajectories of annual MMR, skilled birth attendance (SBA), and contraceptive prevalence rate (CPR), from 2005 to 2023. The exposure was the volume of health-related UPR recommendations received across three cycles, thematically classified using a validated rule-based algorithm. Mixed-effects models adjusted for time-varying GDP per capita and historical fragility.

The 89 countries received 41,733 UPR recommendations across three cycles, of which 405 (1%) were related to maternal health. Maternal health recommendations were preferentially directed at countries with higher baseline MMR and lower SBA. After adjustment, each additional maternal health recommendation was associated with a 0.24% [95% confidence interval (CI): 0.08, 0.40] faster annual reduction in MMR, a 0.52% [0.12, 0.91] faster annual gain in the odds of SBA, and a 0.21% [0.09, 0.34] faster annual gain in the odds of CPR. Broader recommendations on women’s health and health systems and services were also associated with faster annual improvements in trajectories across all three outcomes; recommendations on abortion, family planning, sexual health and wellbeing, and sexual education tended to be directed towards lower-burden countries and were not associated with differences in any trajectories. It is important to note that the ecological design precludes causal inference.

**Conclusions:** Receiving UPR recommendations on the themes of maternal health, women’s health, and health systems and services are associated with accelerated improvements in maternal health trajectories among high-burden countries. These findings suggest that international human rights accountability mechanisms may have a role in supporting national progress on maternal health.

## Introduction

Despite substantial reductions in maternal mortality over the past two decades, preventable maternal deaths remain unacceptably common, particularly in low- and middle-income countries. The global MMR was estimated to be 197 per 100,000 live births in 2023, and at the current rate of progress is expected to reach 177 by 2030, far higher than the 2030 Sustainable Development Goal (SDG) Target of 70 [1,2]. The burden of maternal mortality is not evenly distributed, as low- and lower-middle income countries and those affected by violent conflict or high levels of institutional and social fragility account for 95% of maternal deaths [1,3–5]. Most maternal deaths are preventable, and structural approaches that address their political, economic, and social determinants are needed to accelerate progress [6–8].

Most (173) UN member states are party to the International Covenant on Economic, Social and Cultural Rights, which asserts under Article 12 “the right of everyone to the enjoyment of the highest attainable standard of physical and mental health” [9]. Preventable maternal mortality has been increasingly conceptualized as a violation of this right to health [10]. Accountability – the obligation of States to *monitor* results and resources, *review* progress, and *act* to accelerate progress and remedy failures – is a fundamental pillar for ensuring that health entitlements lead to tangible health improvements [11,12]. One tool to operationalize accountability of all UN member states in their human rights record, including the right to health, is the Universal Periodic Review (UPR). It is a state-driven peer-review mechanism, and three cycles are completed (2008–11, 2012–16, 2017–22) with the fourth one currently ongoing (2022-27) [13,14]. Recommendations are issued by peer (recommending) States, and the State under review can “support” (accept) or “note” (reject) them, with supported recommendations constituting voluntary political commitments to action.

Several plausible pathways through which engagement with the UPR could influence domestic health outcomes have previously been proposed [14–18]. The State under review’s cooperative engagement with the UPR recommendations along with ongoing dialogue and repeated interactions with the recommending States can help shift national attitudes and policies toward human rights compliance over time [15,16]. Supported recommendations from the UPR provide a foothold that civil society organizations and national human rights institutions can use to demand domestic accountability [14,16,18]. As part of the mandatory national reporting phase, State governments are supposed to consult with national stakeholders, offering further avenues for ongoing domestic dialogue and influence. The cyclical nature of the UPR, with a growing focus on reporting on the implementation of recommendations from previous cycles, allows for collaboration and interaction between and across several sectors and ministries, furthering the political positioning of health in the country.

Prior research has catalogued and thematically characterized the rise in health-related recommendations across the first three cycles of the UPR, and has variously described implementation rates [14,19,20]. While prior research has also assessed whether the ratification status of various human rights treaties is associated with real-world health outcomes [21], to our knowledge empirical evidence linking UPR recommendations to measurable improvements in maternal health is not available. We examined whether receiving UPR recommendations during the first three cycles is associated with an accelerated decline in maternal mortality between 2005 and 2023. Reductions in maternal mortality can be driven by improvements in maternity care as well as decreases in the fertility rate [6,22]. To capture these components this study additionally examined the trajectories of two important indicators: the proportion of births attended by skilled health personnel, and the contraceptive prevalence rate. In doing so, this article offers a quantitative assessment of whether receiving UPR recommendations is associated with real-world improvements in maternal health outcomes.

## Methods

### Study design and outcomes

Using a longitudinal ecological analysis, this study examined whether States receiving a higher number of recommendations related to various health-related themes was associated with improved trajectories of the following three outcomes related to maternal health between 2005 and 2023: 1) annual estimates of the Maternal Mortality Ratio (MMR; expressed as the number of maternal deaths per 100,000 live births); 2) annual estimates of the percentage of births attended by skilled health personnel (hereafter referred to as “Skilled Birth Attendance”; SBA); and 3) contraceptive prevalence rate (CPR) defined as the “percentage of women aged 15 to 49 who are currently using, or whose sexual partner is using, at least one method of contraception, regardless of the method used.” MMR and SBA were obtained from WHO’s Global Health Observatory [23]. Estimates of MMR were available for each country for each year between 2005 and 2023, while for SBA the data availability per country varied each year. Annual estimates of CPR were obtained from the UNFPA Population Data Portal [24] and were available between 2005 and 2023 for all States except for Micronesia. The analysis was restricted to UN Member States (hereafter “States”) with an MMR ≥ 70 in 2005 (n = 94), thereby excluding those States that had already achieved the 2030 SDG Target at the start of the study period.

### Exposures

The exposures of interest were the total number of UPR recommendations received across several health-related thematic areas, extracted for the first three cycles of the UPR from the Universal Human Rights Index database [25]. Thematic groupings were adapted from a 2019 WHO review of the first two cycles of the UPR [19], and a rule-based algorithm using keywords was developed to identify, and thematically classify, health-related recommendations from the UPR. A random sample of nearly 15,000 recommendations was manually classified independently by two reviewers (JS and MP), and this was used to validate the rule-based algorithm. Further details regarding the methodology and validation are provided in the supplementary material.

### Statistical Analysis

Descriptive analysis

The baseline and change scores for each of the three outcomes of interest – MMR, SBA, and CPR – were summarized by categorizations of the volume of UPR recommendations received across various health-related themes as well as by several country-level contextual factors: WHO region, World Bank income group in 2005, total years appearing on the World Bank’s list of countries in Fragile and Conflict-affected Situations (FCS), and the baseline MMR in 2005. Change scores for MMR and CPR were calculated as the absolute difference between estimates from 2005 and 2023, while SBA change scores were calculated from the earliest available baseline (minimum year: 2005) until the latest available measure per country (countries with fewer than 5 years between the earliest and latest SBA measures were excluded).

Mixed-effects models with longitudinal data

To evaluate the longitudinal association between receiving UPR recommendations and national trajectories in our outcomes of interest, a series of mixed-effects models were estimated using R. Intercepts and slopes were allowed to vary by country, and the time variable was calculated as years from baseline (2005 = 0) to establish a meaningful intercept. Because the three outcome variables had different underlying distributions, two distinct modeling strategies were used.

Linear mixed-effects models were used to calculate trajectories of MMR, using annual MMR estimates that were natural log-transformed to satisfy the assumption of normally distributed residuals, correct for right skewness, and prevent the estimation of theoretically impossible negative ratios. Fixed-effect coefficients for these models were exponentiated and are interpreted as relative percentage changes.

For SBA and CPR, the percentages were scaled to continuous proportions bounded between 0 and 1, and mixed-effects Beta regression models with logit links were fitted to prevent out-of-bounds predictions. Fixed-effect coefficients for the Beta models were exponentiated and are interpreted as relative percentage changes in the odds of the outcome.

Across all three models, the primary predictor – the time-invariant continuous total count of UPR recommendations received across the first three cycles (for each thematic grouping) – was included as a main effect to adjust for baseline differences, and as a cross-level interaction with time to assess its impact on the annual rate of change. The adjusted models included the following covariates: scaled and centered annual GDP per capita, adjusted for Purchasing Power Parity (PPP) was included as a time-varying control for socioeconomic fluctuations (data from the World Bank’s Open Data portal), and the categorized count of years that a country was listed on the World Bank’s annual classification of Fragile and Conflict-Affected Situations (FCS) was included as a time-invariant measure of fragility. The FCS classifications cover the years 2006 to 2023, and were categorized as 0, 1-8 years, 9-15 years, and 16+ years (the cutoffs for the latter three groups were chosen to obtain approximately equal group sizes). Missing values were not imputed, and complete-case analyses were used throughout.

Data preparation, statistical analyses, and visualization of results were performed using R, version 4.5.0 (https://www.R-project.org/). Mixed-effects models were analyzed using the *lme4* (1.1-37) and *glmmTMB* (1.1.14) packages.

## Results

Of 94 countries with a baseline MMR of 70 or greater in 2005, 5 countries (North Korea, Eritrea, South Sudan, Venezuela, and Yemen) were excluded due to no available timepoints for GDP per capita (PPP) between 2005 and 2023 leaving a total number of 89 countries(Figure S1). For the SBA and CPR analyses, outcome data was not available for Equatorial Guinea and Micronesia, respectively, during the 2005 to 2023 observation period.

Nearly half [48% (43/89)] of the countries were from the WHO African Region (Table 1), and these countries also had the highest median MMR at baseline (median: 467; interquartile range (IQR): 344 to 629). 48 countries (55%) were categorized as low-income in 2005, and these countries had the highest median MMR and lowest median SBA and CPR at baseline. 45 countries (51%) appeared at least once on the Fragile and Conflict-Affected Situations list between 2006 and 2023, and these countries had higher median MMR and lower median SBA and CPR at baseline relative to countries with 0 years on the list.

**Table 1.** Contextual factors by baseline and change scores for each outcome.

|  | Countries<br>N (%)<sup>1</sup> | MMR<br>Median (IQR) |  | SBA<br>Median (IQR) |  | CPR<br>Median (IQR) |  |
| --- | --- | --- | --- | --- | --- | --- | --- |
| Characteristic | N = 89 | Baseline | Change | Baseline | Change | Baseline | Change |
| **WHO region** |  |  |  |  |  |  |  |
| African Region (AFR) | 43 (48%) | 467 (344, 629) | -179 (-293, -74) | 52 (42, 69) | 20 (11, 40) | 19 (12, 29) | 8 (3, 14) |
| Eastern Mediterranean Region (EMR) | 7 (7.9%) | 315 (196, 1,086) | -160 (-523, -126) | 49 (31, 78) | 23 (9, 34) | 11 (6, 29) | 7 (1, 8) |
| South-East Asian Region (SEAR) | 6 (6.7%) | 328 (277, 378) | -200 (-263, -139) | 38 (20, 52) | 45 (40, 50) | 29 (23, 42) | 9 (8, 11) |
| Western Pacific Region (WPR) | 16 (18%) | 175 (105, 296) | -57 (-104, -26) | 86 (62, 96) | 6 (1, 28) | 25 (21, 33) | 2 (-1, 6) |
| Region of the Americas (AMR) | 16 (18%) | 103 (82, 180) | -34 (-78, -14) | 88 (69, 97) | 9 (2, 17) | 41 (35, 50) | 7 (4, 8) |
| European Region (EUR) | 1 (1.1%) | 74 (74, 74) | -32 (-32, -32) | 98 (98, 98) | 2 (2, 2) | 29 (29, 29) | -3 (-3, -3) |
| **WB Income Group, 2005** |  |  |  |  |  |  |  |
| Low income | 48 (55%) | 462 (325, 667) | -190 (-320, -96) | 46 (33, 55) | 30 (16, 43) | 19 (11, 25) | 8 (5, 14) |
| Lower middle income | 32 (37%) | 140 (90, 255) | -59 (-127, -26) | 79 (66, 91) | 10 (3, 24) | 37 (28, 44) | 4 (1, 9) |
| Upper middle income | 6 (6.9%) | 171 (105, 279) | -22 (-138, -11) | 94 (94, 100) | 3 (0, 6) | 36 (26, 51) | 4 (-1, 7) |
| High income | 1 (1.1%) | 70 (70, 70) | 6 (6, 6) | 99 (99, 99) | 0 (0, 0) | 38 (38, 38) | 7 (7, 7) |
| Missing | 2 |  |  |  |  |  |  |
| **Years on FCS list, 2006-2023** |  |  |  |  |  |  |  |
| 0 | 44 (49%) | 190 (97, 336) | -62 (-148, -26) | 78 (52, 95) | 13 (3, 38) | 36 (27, 44) | 7 (2, 9) |
| 1-8 | 18 (20%) | 460 (283, 629) | -192 (-346, -95) | 51 (34, 59) | 17 (9, 42) | 17 (10, 26) | 9 (4, 16) |
| 9-15 | 12 (13%) | 356 (209, 535) | -109 (-177, -60) | 55 (52, 86) | 17 (7, 29) | 21 (11, 29) | 5 (2, 9) |
| 16+ | 15 (17%) | 619 (373, 1,086) | -249 (-456, -96) | 41 (29, 64) | 23 (15, 34) | 15 (10, 21) | 8 (3, 10) |
| **Abortion laws (2024)** |  |  |  |  |  |  |  |
| I. On Request | 13 (15%) | 279 (99, 323) | -106 (-186, -35) | 80 (50, 96) | 10 (3, 21) | 28 (16, 42) | 6 (1, 11) |
| II. Socioeconomic Grounds | 4 (4.5%) | 371 (271, 610) | -217 (-398, -188) | 42 (23, 46) | 47 (43, 52) | 19 (11, 35) | 13 (9, 22) |
| III. To Preserve Health | 30 (34%) | 412 (196, 613) | -140 (-279, -52) | 63 (46, 81) | 17 (7, 30) | 23 (14, 38) | 7 (4, 9) |
| IV. To Save the Mother's Life | 26 (29%) | 298 (186, 505) | -128 (-247, -59) | 53 (42, 86) | 24 (6, 39) | 23 (14, 34) | 6 (3, 9) |
| V. Prohibited Altogether | 16 (18%) | 163 (99, 473) | -58 (-150, -26) | 73 (48, 93) | 13 (3, 28) | 28 (20, 39) | 6 (2, 10) |
| **MMR category, 2005** |  |  |  |  |  |  |  |
| 70-119 | 19 (21%) | 87 (75, 103) | -25 (-35, -4) | 95 (78, 99) | 4 (1, 19) | 39 (29, 45) | 3 (-1, 8) |
| 120-219 | 13 (15%) | 175 (161, 188) | -64 (-118, -38) | 87 (77, 91) | 10 (2, 16) | 32 (27, 44) | 7 (1, 11) |
| 220-299 | 9 (10%) | 268 (242, 279) | -128 (-179, -79) | 62 (55, 86) | 13 (7, 40) | 35 (27, 41) | 5 (-1, 10) |
| 300+ | 48 (54%) | 490 (376, 667) | -204 (-351, -130) | 46 (32, 57) | 26 (13, 42) | 17 (10, 24) | 8 (4, 13) |

<sup>1</sup>% may not add up to 100% due to rounding
IQR: Interquartile range

Across the first three cycles of the UPR, the 89 countries from the analytical sample received a total of 41,733 UPR recommendations (459; IQR: 386 to 552), of which 9,852 (24%) were classified as health-related (113; IQR: 84 to 133). The most commonly identified themes were “Gender-based violence and harmful practices” (71; IQR: 50 to 84), “Child and adolescent health” (30; IQR: 24 to 39), “Health systems and services” (16; IQR: 13 to 22), “Sexual health and wellbeing” (7; IQR: 3 to 14), and “Maternal health” (4; IQR: 2 to 6) (Figure S2).

At baseline (Table 2), the median MMR was 315 per 100,000 live births (IQR: 161 to 505), median SBA coverage was 61% (IQR: 44 to 86), and median CPR was 25% (IQR: 15 to 38). Over the study period, all three indicators moved in the expected direction: MMR declined by a median of 126 deaths per 100,000 live births (IQR: −210 to −39), SBA increased by a median of 17% (IQR: 6% to 37%), and CPR increased by a median of 7% (IQR 2% to 10%).

**Table 2.** UPR recommendations by baseline and change scores for each outcome.

| Characteristic | Countries<br>N (%)<sup>1</sup> | MMR<br>Median (IQR) |  | SBA<br>Median (IQR) |  | CPR<br>Median (IQR) |  |
| --- | --- | --- | --- | --- | --- | --- | --- |
|  | N = 89 | Baseline | Change | Baseline | Change | Baseline | Change |
| **Overall sample:** |  | 315 (161, 505) | -126 (-210, -39) | 61 (44, 86) | 17 (6, 37) | 25 (15, 38) | 7 (2, 10) |
| **UPR recommendations (n)** |  |  |  |  |  |  |  |
| **Health-related (any)** |  |  |  |  |  |  |  |
| 1-94 | 31 (35%) | 202 (105, 323) | -67 (-153, -25) | 79 (51, 94) | 13 (2, 26) | 28 (19, 40) | 5 (2, 9) |
| 95+ | 58 (65%) | 388 (181, 540) | -141 (-263, -58) | 55 (40, 81) | 22 (7, 39) | 23 (13, 37) | 8 (4, 10) |
| **Abortion** |  |  |  |  |  |  |  |
| 0 | 33 (37%) | 373 (188, 685) | -138 (-263, -55) | 50 (32, 89) | 17 (6, 29) | 20 (12, 29) | 5 (2, 8) |
| 1-2 | 42 (47%) | 279 (153, 462) | -122 (-197, -34) | 62 (47, 86) | 13 (3, 39) | 27 (17, 39) | 7 (2, 11) |
| 3+ | 14 (16%) | 263 (103, 488) | -96 (-247, -52) | 64 (44, 79) | 24 (3, 40) | 38 (19, 49) | 9 (6, 16) |
| **Family planning** |  |  |  |  |  |  |  |
| 0 | 30 (34%) | 406 (186, 629) | -138 (-263, -35) | 52 (41, 78) | 17 (7, 34) | 21 (11, 34) | 5 (3, 8) |
| 1-2 | 33 (37%) | 350 (242, 462) | -118 (-194, -55) | 59 (44, 85) | 17 (3, 39) | 25 (18, 33) | 8 (4, 14) |
| 3+ | 26 (29%) | 188 (104, 312) | -99 (-197, -34) | 73 (54, 86) | 17 (3, 29) | 32 (17, 43) | 7 (2, 10) |
| **GBV and harmful practices** |  |  |  |  |  |  |  |
| 1-58 | 32 (36%) | 247 (123, 351) | -95 (-183, -31) | 74 (44, 93) | 16 (2, 42) | 28 (19, 40) | 6 (2, 10) |
| 59+ | 57 (64%) | 385 (166, 540) | -136 (-249, -55) | 56 (44, 81) | 17 (7, 33) | 23 (13, 37) | 8 (4, 10) |
| **Health systems and services** |  |  |  |  |  |  |  |
| 1-15 | 36 (40%) | 178 (93, 384) | -57 (-156, -25) | 81 (53, 96) | 7 (2, 19) | 28 (18, 39) | 3 (0, 9) |
| 16+ | 53 (60%) | 373 (268, 529) | -144 (-247, -71) | 52 (40, 73) | 24 (11, 42) | 23 (14, 35) | 8 (5, 11) |
| **Maternal health** |  |  |  |  |  |  |  |
| 0 | 4 (4.5%) | 133 (89, 369) | -32 (-124, -21) | 76 (58, 93) | 9 (-1, 23) | 25 (16, 34) | 5 (1, 9) |
| 1-3 | 35 (39%) | 315 (118, 536) | -67 (-202, -32) | 58 (40, 93) | 12 (3, 29) | 23 (13, 33) | 5 (2, 8) |
| 4+ | 50 (56%) | 335 (186, 492) | -135 (-236, -64) | 59 (45, 81) | 22 (8, 42) | 26 (16, 41) | 8 (3, 11) |
| **Sexual education** |  |  |  |  |  |  |  |
| 0 | 40 (45%) | 338 (181, 624) | -138 (-264, -44) | 55 (43, 83) | 17 (6, 34) | 23 (11, 34) | 6 (2, 10) |
| 1-2 | 33 (37%) | 355 (124, 467) | -96 (-225, -39) | 55 (39, 79) | 17 (7, 42) | 26 (16, 39) | 8 (5, 11) |
| 3+ | 16 (18%) | 188 (129, 304) | -99 (-149, -45) | 81 (67, 91) | 11 (2, 27) | 29 (23, 43) | 5 (0, 10) |
| **Sexual health and wellbeing** |  |  |  |  |  |  |  |
| 0 | 4 (4.5%) | 244 (125, 445) | -81 (-169, 12) | 71 (57, 84) | 12 (1, 18) | 28 (15, 38) | 7 (-3, 11) |
| 1-6 | 39 (44%) | 315 (105, 514) | -133 (-236, -27) | 65 (33, 92) | 14 (3, 30) | 24 (13, 35) | 7 (3, 9) |
| 7+ | 46 (52%) | 327 (181, 488) | -116 (-225, -55) | 57 (46, 81) | 21 (8, 42) | 25 (17, 39) | 8 (2, 11) |
| **Women's health** |  |  |  |  |  |  |  |
| 0 | 10 (11%) | 102 (82, 166) | -33 (-59, -23) | 97 (91, 99) | 2 (1, 7) | 29 (26, 38) | 2 (0, 7) |
| 1-3 | 47 (53%) | 312 (161, 514) | -126 (-247, -52) | 65 (42, 87) | 16 (4, 29) | 27 (16, 39) | 5 (2, 9) |
| 4+ | 32 (36%) | 420 (278, 563) | -157 (-238, -65) | 52 (40, 69) | 29 (11, 42) | 20 (12, 33) | 8 (5, 15) |

<sup>1</sup>n (%) ; % may not add up to 100% due to rounding
IQR: Interquartile range
GBV: Gender-based violence

Countries that received more health-related UPR recommendations had higher median MMR and lower median SBA and CPR at baseline (Table 2), and this pattern largely held true for each of the thematic groupings of UPR recommendations. However, for the themes of abortion, family planning, and sexual education, countries that received more recommendations had lower median MMR and higher SBA and CPR at baseline.

Countries with higher MMR at baseline tended to have larger absolute declines in MMR, and a similar gradient was observed for SBA and CPR, where countries with lower baseline values had larger absolute gains.

### Mixed-effects models with longitudinal data

Univariable (unadjusted) and multivariable (adjusted) estimates for all thematic groupings are presented in Figure 1.

**Figure 1.**
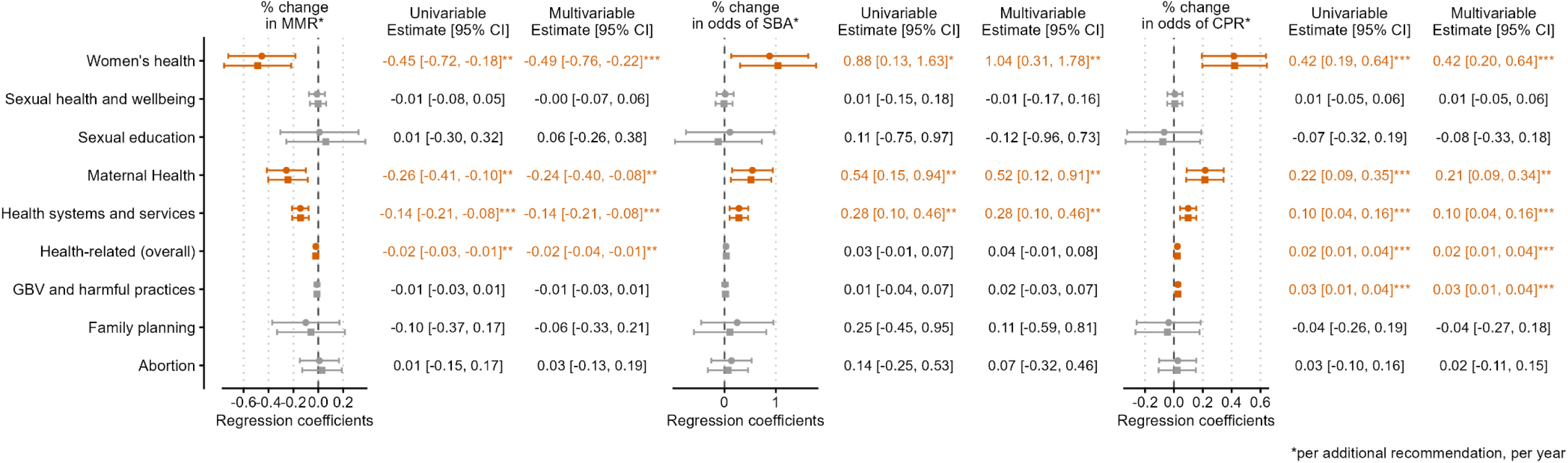
Results of the mixed-effects models for the associations between UPR recommendation volume and longitudinal trajectories for three outcomes: MMR (maternal mortality ratio), SBA (skilled birth attendance), and CPR (contraceptive prevalence rate). The estimates displayed are from the interaction term (UPR recommendations * Year) and represent the estimated annual rate of change per additional recommendation, within each thematic grouping of the UPR recommendations. Univariable models included as main effects the number of recommendations received over the first three cycles of the UPR, Year (baseline in 2005 set to 0) and the interaction of “Year x Number of recommendations”. Slopes and intercepts were allowed to vary by country. The multivariable models additionally included as covariates the scaled and grand-mean centered annual estimates of GDP per capita (PPP), as well as a categorization of the total number of years that a country was listed on the Fragile and Conflict-Affected Situations list (2006 to 2023). For each thematic grouping, the point estimates and 95% confidence intervals of the outcome trajectories are graphically displayed as forestplots, with numerical estimates also displayed in separate columns. Estimates in red and asterisks denote statistical significance (p < 0.05* ; p < 0.01** ; p < 0.001***).

UPR recommendations related to maternal health

Adjusting for covariates, the models demonstrated significant underlying global improvements across all three outcome measures of the study. A significant and consistent association was observed between the number of maternal health related recommendations and improvements in the rates of change for each outcome: each additional recommendation related to maternal health was associated with a −0.24% (95% CI [-0.40%, −0.08%]) annual reduction in MMR, a 0.52% (95% CI [0.12%, 0.91%]) annual increase in the odds of SBA, and a 0.21% (95% CI [0.09%, 0.34%]) annual increase in the odds of CPR.

Across all three outcome measures, negative correlations between countries’ baseline estimates and rates of change showed that countries with worse estimates at baseline tended to show the most rapid improvements over time. Additionally, countries that received a greater number of maternal health recommendations had higher baseline MMR (+6.9% per additional recommendation; p = 0.006) and lower baseline odds of SBA (−10.2% per additional recommendation, p=0.027), but no significant differences were observed for the baseline odds of CPR (+1.08% per additional recommendation; p=0.609).

Other thematic groupings of the UPR recommendations

Broader recommendations related to “Women’s health” (not related to maternal health or other SRHR themes, and including recommendations related to, for example, menstrual health and women’s access to general health services) and “Health systems and services” (including recommendations addressing the broader health system and the population’s general access to health services) were also associated with accelerated improvement across all three outcome measures. Recommendations related to Women’s health yielded the largest effect sizes observed in the analysis: each additional recommendation was associated with a −0.49% (95% CI [-0.76%, −0.22%]) annual reduction in MMR, a 1.04% (95% CI [0.31%, 1.78%]) annual increase in the odds of SBA and a 0.42% (95% CI [0.20%, 0.64%]) annual increase in the odds of CPR. Similarly, recommendations related to “Health systems and services” were associated with faster annual improvements for MMR (−0.14%; 95% CI [-0.21%, −0.08%]), SBA (0.28%; 95% CI [0.10%, 0.46%]), and CPR (0.10%; 95% CI [0.04%, 0.16%]). Recommendations related to “Gender-based violence and harmful practices” were associated with a 0.03% (95% CI [0.01%, 0.04%]) annual increase in the odds of CPR, but were not associated with the rates of change for MMR or SBA. Recommendations related to abortion, family planning, sexual health and wellbeing, and sexual education, were not associated with any differences in the rates of change for any of the three outcomes.

## Discussion

To our knowledge, this is the first study to provide quantitative evidence that receiving UPR recommendations is associated with improved maternal health outcomes. Across 89 high-burden countries, recommending States tended to direct maternal health-related recommendations towards countries with the highest baseline MMR and lowest SBA coverage. After adjusting for GDP per capita at PPP and historical fragility, receipt of more maternal health-related recommendations was associated with faster annual reductions in MMR and faster gains in SBA and CPR between 2005 and 2023. Aside from the recommendations directly related to maternal health, those related to “health systems and services” and “women’s health”, were also associated with improvement in all three indicators, suggesting that recommendations addressing structural determinants and health system issues have meaningful impact on the ground.

Critics have argued that the UPR often operates more as a hollow ritual than as an effective accountability mechanism [26]. Since recommendations are politically negotiated, non-binding, weakly enforced, and often lack specificity, States may appear to engage with international scrutiny without committing to measurable legal, budgetary, or health-system reforms [14,26,27]. Our findings provide empirical evidence to challenge this view, and the value of the UPR should also be considered within the broader context in which the right to health is operationalized. WHO has played an important role in developing the normative basis for recognizing health as a fundamental human right [28]. However, recent global health law reforms, including the 2024 amendments to the International Health Regulations and the negotiation of the WHO Pandemic Agreement, have reduced human rights to largely guiding principles rather than binding legal obligations [29]. The need to reach consensus among Member States limits the strength of human rights commitments achievable through the WHO governance mechanisms alone [28]. The UPR may offer a complementary accountability mechanism within the international human rights architecture, with broad universal coverage across each UN Member State. In the first three cycles every member of the UN participated in the UPR process.

The UPR’s cyclical structure facilitates repeated interactions with peer countries, and this ongoing dialogue and periodic reporting helps to gradually achieve normative change within the State under review [15,16,30]. The pattern observed in our data, where additional recommendations were associated with incrementally faster improvement, is consistent with this framework. Sustained engagement by peer countries may encourage States to strengthen health-related commitments with each review cycle progressively. However, the impact of recommendations may depend in part on the normative consensus on the issues raised. While maternal health, health systems improvement, and gender equity attract broad political support, we found recommendations related to SRHR (abortion, family planning, sexual health and wellbeing, and sexual education) were not associated with any differences in trajectories for any of the three outcomes. This may partly reflect a selection bias. Unlike maternal health recommendations, which were directed at high-burden States, these recommendations tended to target countries with better baseline health indicators. As a result, there was less observable improvement in our models. This observation may also be attributed to limited normative consensus around the themes. Consistent with previous findings [14], our data reveal that these recommendations tend to be issued by only a small group of countries, mostly from the European Region. This may reduce the benefit from broader interaction with peer countries.

Reducing maternal mortality at scale ultimately depends on health system strengthening and progress toward universal health coverage [6], neither of which can be sustained without political commitment. The UPR may help reinforce this by generating and regularly reviewing the documented political commitments against which progress can be measured. Health-related recommendations may be particularly well-suited to this mechanism, as they tend to enjoy broader normative consensus and higher acceptance rates than recommendations on more politically contested themes, such as civil and political rights [14]. The resulting volume of accepted health-related commitments offers civil society organizations, national human rights institutions, and parliamentary stakeholders a concrete entry point for advocacy using documented State commitments to press for domestic resource allocation, policy reform, and accountability for implementation [16,18].

The relevance of this dimension of domestic accountability has grown in light of ongoing cuts to global health financing. As external financing recedes, governments face complex choices about how to sustain critical services, some of which carry serious risks for equity, including reverting to out-of-pocket payments, over-reliance on contributory insurance, or abandoning community-based delivery models [31]. In this context, mechanisms that support domestic political accountability for equitable health investment become particularly important. The UPR provides a forum through which States can publicly commit to health priorities, including through voluntary pledges that carry into subsequent reviews, an under-utilized pathway for signaling domestic ownership of health at a moment when such ownership matters most.

This study has several strengths. It is, to our knowledge, the first to empirically link receiving UPR recommendations to downstream health outcomes. The analysis spans 18 years and three complete UPR cycles, and the findings are consistent across three distinct indicators of maternal health. The text classification algorithm allowed for a reproducible coding of the UPR recommendations, and this was validated against a manually classified sample of nearly 15,000 recommendations.

Several limitations should be considered when interpreting these findings. This study uses an ecological design, and associations between recommendation volume and national health trajectories cannot be attributed to changes experienced by specific populations within those countries [32]. Observed associations could reflect unmeasured confounding (which could independently drive both UPR engagement and health improvement. Similarly, the associations could reflect reverse causation, whereby recommending States are more likely to issue recommendations to countries already committed to improving maternal health. The faster improvement we observe among countries with worse baseline estimates is consistent with the broader pattern of health convergence, where high-burden countries have the greatest scope for improvement and tend to advance most rapidly as proven interventions are scaled up [3,33]. Our mixed-effects framework partially accounts for this by giving each country its own baseline and rate of change, so country-specific differences in underlying trajectories are absorbed into the random effects rather than attributed to UPR engagement. Treating recommendation volume as a time-invariant exposure limits our ability to capture the temporal sequence of health diplomacy versus health improvements. We do not account for recommendation quality or whether they were accepted or merely noted; however, even when recommendations are formally noted they may still drive domestic engagement and action [17], and so it was deemed appropriate to use all recommendations for the analysis. Future studies could account for the number of States attached to each recommendation. Near-identical recommendations from multiple States are typically merged into a single entry, so our analysis does not distinguish between a recommendation issued by one State and one issued by many, which may itself reflect the strength of normative consensus.

Despite these limitations, this study suggests that the UPR is not a purely rhetorical exercise; it is associated with measurable accelerations in maternal health progress among the countries that need it most. For States formulating future recommendations, our results suggest that broadly framed recommendations addressing structural determinants and health systems may be as important as those targeting maternal health directly. While we cannot test this directly, broader engagement by peer States on contested SRHR themes (currently issued by only a small group of States) may, over successive review cycles, help build the normative consensus on which the UPR’s accountability function depends.

## Data Availability

Data is available at https://github.com/CeHDI-Foundation/UPR_Maternal-Health_Public

## Supplementary material

### Identification of health-related recommendations and thematic classification

A rule-based text classification algorithm was developed to identify health-related themes from all available UPR recommendations (obtained from the Universal Human Rights Index), which were thematically classified using groupings adapted from those developed in a 2019 report from WHO [19]. For each theme, a dictionary of keywords and term combinations was constructed and then matched against the recommendation text to determine theme correspondence. A single recommendation could match several themes.

In the example of the theme of “Maternal health” the dictionary of keywords included terms such as “obstetric,” “prenatal,” “postnatal,” “miscarriage,” and “maternal mortality.” The algorithm also identified recommendations containing specific combinations of terms linking pregnancy to healthcare access (e.g., “pregnant” appearing in conjunction with “healthcare,” “medical care,” or “free access”). Recommendations primarily addressing abortion were conditionally classified as maternal health only if they explicitly contextualized abortion access within the framework of saving the woman’s life or preserving her physical health. Linguistic false positives (e.g., “miscarriage of justice”) were explicitly excluded for each theme. Similar steps were followed when elaborating the dictionary of keywords for each of the thematic groupings.

To ensure reproducibility, these recommendation classifications were coded using R. The full code and datasets are available on GitHub (https://github.com/CeHDI-Foundation/UPR_Maternal-Health_Public) and the codebook of operational definitions that was used to guide the coding are included in this supplementary material below.

Validation of thematic classifications

A two-reviewer manual classification of a random subset of 14,700 of all available recommendations (∼12% of the ∼123,000 recommendations extracted from the Universal Human Rights Index) was undertaken to assess the algorithm’s performance. After familiarizing themselves with the codebook of operational definitions, the two reviewers independently classified each of the randomly selected UPR recommendations into one or more of the thematic groupings. Recommendations that were deemed to be not related to health were marked “Not health-related”. For all classifications where no discrepancies occurred between the reviewers, a gold-standard “reference” classification was extracted in a separate column. In case of discrepancies the reviewers agreed on the final classification, and these values were entered into the reference column.

For each thematic grouping, Cohen’s kappa was used to assess agreement between the two reviewers. The reference values were used to calculate the sensitivity and positive predictive value (PPV) for each of the thematic groupings identified by the automated classifications.

The automated classification demonstrated high reliability and validity for most of the thematic groupings (Table S1). For the “Maternal health” thematic grouping, the automated algorithm achieved a sensitivity of 0.855 and a positive predictive value (PPV) of 0.969. The classifications for the themes of “Abortion”, “Sexual education”, and “Sexual health and wellbeing” were similarly robust. However, findings were less robust for “Women’s health” (sensitivity: 0.8, PPV: 0.693), “Health systems and services” (sensitivity: 0.768, PPV: 0.685), and “Family planning” (sensitivity: 0.974, PPV: 0.826).

**Table S1.** Validation metrics to assess the automatic and manual classifications.

| UPR Theme | Reviewer 1<br>(n) | Reviewer 2<br>(n) | Kappa | Reference<br>(n) | Algorithm<br>(n) | Sensitivity | PPV |
| --- | --- | --- | --- | --- | --- | --- | --- |
| **Abortion** | **71** | **69** | **0.928** | **73** | **73** | **1.000** | **1.000** |
| Child and adolescent health | 756 | 762 | 0.843 | 812 | 827 | 0.931 | 0.913 |
| Communicable diseases | 20 | 9 | 0.551 | 13 | 13 | 0.923 | 0.923 |
| Disabilities and health | 60 | 68 | 0.718 | 68 | 71 | 0.868 | 0.831 |
| **Family planning** | **29** | **41** | **0.771** | **39** | **46** | **0.974** | **0.826** |
| GBV and harmful practices | 1,849 | 1,817 | 0.943 | 1,880 | 1,848 | 0.971 | 0.987 |
| HIV/AIDS and STIs | 56 | 54 | 0.945 | 56 | 58 | 1.000 | 0.966 |
| Health of LGBTQI persons | 21 | 9 | 0.133 | 15 | 8 | 0.467 | 0.875 |
| Health of incarcerated persons | 53 | 51 | 0.768 | 53 | 61 | 0.868 | 0.754 |
| Health emergencies | 45 | 40 | 0.445 | 51 | 32 | 0.392 | 0.625 |
| **Health systems and services** | **406** | **397** | **0.773** | **449** | **504** | **0.768** | **0.685** |
| Health-related | 3,014 | 2,930 | 0.927 | 3,036 | 2,991 | 0.962 | 0.976 |
| **Maternal health** | **138** | **134** | **0.896** | **145** | **128** | **0.855** | **0.969** |
| Mental health | 74 | 79 | 0.744 | 87 | 65 | 0.678 | 0.908 |
| Non-communicable diseases | 12 | 8 | 0.500 | 13 | 10 | 0.769 | 1.000 |
| Nutrition | 85 | 78 | 0.846 | 90 | 61 | 0.656 | 0.967 |
| Other health-related | 83 | 30 | 0.317 | 51 | 79 | 0.392 | 0.253 |
| Refugee and migrant health | 59 | 39 | 0.591 | 50 | 77 | 0.900 | 0.584 |
| **Sexual education** | **44** | **44** | **0.818** | **54** | **50** | **0.907** | **0.980** |
| **Sexual health and wellbeing** | **234** | **239** | **0.903** | **237** | **237** | **0.949** | **0.949** |
| TB, malaria, and NTDs | 14 | 9 | 0.782 | 14 | 14 | 0.929 | 0.929 |
| Water and sanitation | 88 | 81 | 0.875 | 89 | 71 | 0.798 | 1.000 |
| **Women's health** | **71** | **67** | **0.680** | **65** | **75** | **0.800** | **0.693** |

Random sample for classification performance: N = 14700
Reference: 'Gold-standard' values created after resolving differences in the manual classifications
Algorithm: Values generated from the automated classifications
PPV: Positive predictive value

### Codebook of operational definitions for thematic classification of health-related UPR recommendations

This codebook outlines the operational definitions used to classify health-related recommendations issued under the Universal Periodic Review (UPR) into thematic categories. The framework was adapted from a 2019 World Health Organization (WHO) review of the first two cycles of the UPR [19] and supplemented, where relevant, with definitions drawn from WHO health topic pages (https://www.who.int/health-topics). Definitions were paraphrased and refined through iterative team discussion, and decision rules were added to handle overlapping or ambiguous cases. Each category is presented below with its operational definition, illustrative examples, and any relevant exclusions or cross-classification rules.

This codebook was used to guide the automated identification and classification of UPR recommendations related to health.

Health systems and services

This category captures recommendations addressing the broader health system and the population’s general access to health services. It draws on WHO’s framework of six health system building blocks: service delivery, health workforce, health information systems, access to essential medicines and technologies, health financing, and leadership and governance. Recommendations relating to immunization programs, vaccination campaigns, and the general availability of medicines and pharmaceutical products are also included here.

Illustrative examples include recommendations on the training and deployment of healthcare workers, the implementation of universal health coverage, increases in domestic health financing, the protection of the right to health for the general population, and the expansion of general health services.

**Decision rule:** This category is reserved for recommendations addressing the general population and the broader health system. Recommendations targeting specific populations or issues are classified under their respective categories instead (e.g. recommendations on improving SRHR services are classified under the relevant SRHR sub-category, and recommendations on expanding services for women are classified under Women’s health). When a recommendation simultaneously calls for general system-level improvements alongside actions on a specific issue or population, it is dual-classified under Health systems and services and the relevant sub-category.

Health security, emergencies, and disaster relief

This category covers recommendations addressing health in the context of humanitarian emergencies, disasters, and other situations posing significant threats to population health. Such situations include outbreaks of disease, exposure to contaminated environments, and large-scale events (whether natural or human-made) that affect the health and well-being of substantial population groups and require coordinated multi-sectoral responses.

Illustrative examples include recommendations relating to pandemic preparedness and response (e.g. COVID-19), humanitarian assistance contributing to health outcomes, and relief efforts following natural disasters such as earthquakes, tsunamis, or hurricanes.

**Decision rule:** Recommendations addressing strictly non-health dimensions of emergencies (e.g. the economic consequences of a pandemic) are excluded from this category.

Non-communicable diseases

This category encompasses recommendations addressing non-communicable diseases (NCDs), including cardiovascular disease, cancer, chronic respiratory conditions (such as chronic obstructive pulmonary disease and asthma), diabetes, and substance use disorders involving alcohol and drugs. Other NCDs not explicitly listed are also included.

Illustrative examples include recommendations on tobacco control, the regulation of alcohol consumption, treatment for substance use, and the regulation of air pollution as a determinant of chronic disease.

Communicable diseases

This category includes recommendations addressing infectious and zoonotic diseases. Infectious diseases are those caused by pathogenic microorganisms – including bacteria, viruses, parasites, and fungi – that can be transmitted directly or indirectly between people. Zoonotic diseases are infectious diseases of animals that can be transmitted to humans.

Illustrative examples include recommendations calling for increased allocation of resources to curb the spread of infectious diseases.

**Decision rule:** Recommendations relating to COVID-19 are excluded from this category and instead classified under Health security, emergencies, and disaster relief. Recommendations related to HIV, STIs, tuberculosis and malaria and other Neglected Tropical Diseases are excluded from this category and instead classified under “HIV/AIDS and STIs” and “TB, Malaria, and NTDs”.

Sexual health and wellbeing

This category captures recommendations addressing sexual health and wellbeing as defined by WHO: a state of physical, mental, and social well-being in relation to sexuality, grounded in a positive and respectful approach to sexuality and free from coercion, discrimination, and violence. The category also includes recommendations on infertility and on the recognition of sexual rights more broadly.

Illustrative examples include recommendations on sexual health services, the decriminalization of consensual sexual activity, the decriminalization of sexual orientation or gender identity, and recommendations relating to voluntary sex work.

**Decision rule:** This category does not include recommendations specifically addressing maternal health, abortion, family planning, contraception, or comprehensive sexuality education, which are classified under their respective categories.

Sexual education

This category captures recommendations addressing comprehensive sexuality education, including curriculum content, age-appropriate delivery, and the integration of sexual and reproductive health information into formal and non-formal educational settings.

Illustrative examples include recommendations to introduce or expand comprehensive sexuality education in schools, to ensure age-appropriate sexual health information for adolescents, and to train educators on sexuality education.

Mental health

This category covers recommendations addressing mental health, conceptualized broadly to encompass the promotion of mental well-being, the prevention of mental disorders, and the treatment and rehabilitation of people affected by mental health conditions.

Recommendations addressing mental disabilities, suicide and suicide prevention, and psychosocial development or support services are included in this category.

Gender-based violence and harmful practices

This category covers recommendations addressing violence perpetrated against individuals on the basis of sex, gender identity or expression, or sexual orientation, as well as harmful traditional practices. Harmful practices include - but are not limited to - female genital mutilation, female infanticide, sex-selective abortion, conversion therapy, forced and early marriage, levirate marriage, so-called honour crimes, polygamy, wife inheritance, dowry-related violence, virginity testing, and breast ironing.

Illustrative examples include recommendations on rape, coercive abortion, forced sterilization, domestic and intimate partner violence, sexual violence and abuse, violence against women, forced sex work or sexual exploitation, dowry-related violence, and the inhumane treatment or abuse of LGBTQI individuals. Recommendations using the general phrase “harmful practices” are also classified here.

**Cross-classification rules:** Recommendations on violence, rape, or forced sex work involving children are dual-classified under Child and adolescent health. Recommendations on corporal punishment fall under this category only when they are specifically situated in the context of children, schools, the home, or “all settings”, in which case they are also dual-classified under Child and adolescent health. Recommendations referencing the Istanbul Convention are classified under this category. Child and early marriage is classified under this category alone (not under Child and adolescent health). Sexual trafficking, exploitation, and harassment of women and girls are classified under this category, with dual classification under Child and adolescent health when girls or boys are explicitly mentioned. Forced abortion is dual-classified with Abortion. Early, adolescent, or teenage pregnancy is classified under this category when it occurs in the context of violence; abortion in the context of rape or incest is dual-classified with Abortion.

Women’s health

This category captures recommendations addressing women’s health from a general health-system perspective rather than through the lenses of violence, sexual and reproductive health and rights, maternal health, family planning, abortion, sexual health, or infertility (which are classified under their respective categories). The category captures the application of a gendered lens to general health concerns.

Illustrative examples include recommendations on menstrual health, cervical cancer screening and treatment, and women’s access to general health services.

Child and adolescent health

This category covers recommendations addressing fetal, newborn, infant, child, and adolescent health.

Illustrative examples include recommendations on infant mortality, violence against children (including corporal punishment), child abuse, sexual abuse of children, and childhood cancer. Recommendations on early or teenage pregnancies are dual-classified with Maternal health.

**Decision rule:** Recommendations on early or child marriage are not classified here; they fall under Gender-based violence and harmful practices. Recommendations on corporal punishment are classified under this category (with dual classification under Gender-based violence and harmful practices) only when they are clearly situated in the context of children, schools, the home, or “all settings”; references to corporal punishment without such context are not considered health-related.

Maternal health

This category covers recommendations addressing maternal health, including pregnancy, childbirth, and the postpartum period.

Illustrative examples include recommendations on antenatal and postnatal care, free maternity care, maternal mortality reduction, and obstetric care. Recommendations on early, teenage, or child pregnancies are dual-classified with Child and adolescent health.

**Decision rule:** Recommendations focused exclusively on preventing or reducing early or teenage pregnancies - without addressing care for active pregnancies - are not classified under Maternal health and are instead classified under Family planning and contraception. Recommendations related to abortion are classified only under Abortion.

Abortion

This category covers recommendations addressing abortion, conceptualized within WHO’s framing of comprehensive abortion care as an essential component of universal access to quality health services. Comprehensive abortion care encompasses the provision of information, the management of abortion procedures, and the delivery of post-abortion care.

Illustrative examples include recommendations referring to abortion, the interruption/termination of pregnancies, post-abortion care, and the legalization or decriminalization of abortion.

**Cross-classification rules:** Recommendations addressing forced or coercive abortions are dual-classified with Gender-based violence and harmful practices.

Family planning and contraception

This category covers recommendations addressing family planning and contraception. Access to family planning is grounded in the right of individuals to make informed choices about their sexual and reproductive health.

Illustrative examples include recommendations on access to modern contraceptive methods (e.g. condoms, intrauterine devices, oral contraceptives), emergency contraception, and the prevention of unintended pregnancies, as well as those addressing infertility. This category also includes recommendations using language such as “reducing teenage pregnancies” or “bringing down the rate of early pregnancies”, where the focus is on prevention rather than care during an active pregnancy.

Disabilities and health

This category covers recommendations addressing the health and access to health services of persons with disabilities. “Disability” is understood broadly to include impairments, activity limitations, and participation restrictions.

**Decision rule:** Recommendations not explicitly addressing health issues, e.g, those aimed at curbing discrimination against persons with disabilities outside of a health context, are not included under this category.

Health of LGBTQI persons

This category covers recommendations addressing the health and access to health services of lesbian, gay, bisexual, transgender, queer, and intersex (LGBTQI) persons. It includes recommendations addressing both general health services and LGBTQI-specific health needs, such as gender-affirming care including hormone therapy and gender-affirming surgery.

HIV/AIDS and sexually transmitted infections

This category covers recommendations addressing HIV/AIDS and other sexually transmitted infections (STIs), including access to prevention, testing, treatment, and care for people living with HIV/AIDS or STIs, as well as broader prevention efforts.

Tuberculosis, malaria, and neglected tropical diseases

This category covers recommendations addressing tuberculosis, malaria, and neglected tropical diseases. Neglected tropical diseases include - but are not limited to - rabies, leprosy, foodborne trematodiases, lymphatic filariasis, dracunculiasis (Guinea-worm disease), human African trypanosomiasis (sleeping sickness), and chikungunya.

Water and sanitation

This category covers recommendations addressing access to safe drinking water and basic sanitation services. It includes recommendations on drinking-water quality, water supply and sanitation monitoring, surveillance and prevention of waterborne diseases such as cholera, water and sanitation across different settings, and the management of water resources.

Nutrition

This category covers recommendations addressing nutrition and diet-related health concerns, including obesity, malnutrition, and micronutrient deficiencies.

Recommendations on food are classified here when they relate specifically to nutritional content or supplementation.

Health of incarcerated persons

This category covers recommendations addressing the health of incarcerated persons, grounded in the principle that detained individuals are entitled to the same standard of medical care as the general population. The health of incarcerated persons depends not only on health service provision within places of detention but also on broader institutional conditions and practices.

**Decision rule:** Recommendations are classified here only when they specifically address health-related dimensions (e.g. physical health, mental health, sanitation, nutrition). Recommendations addressing torture or corporal punishment in detention are not classified under this category.

Occupational health

This category covers recommendations addressing the promotion and protection of physical, mental, and social well-being among workers across all occupations. Relevant areas include the maintenance and promotion of workers’ health and capacity to work, improvements to working conditions and the working environment to support occupational safety, and the development of work organization and managerial systems consistent with occupational health and safety.

Illustrative examples include recommendations on unhealthy working conditions and on occupational safety and health more broadly.

Refugee and migrant health

This category covers recommendations addressing the health of refugees, asylum seekers, and migrants.

Illustrative examples include recommendations addressing the right to health and access to health services for refugees, asylum seekers, and migrants.

Other health-related

This category captures recommendations that are clearly health-related but that do not fall under any of the categories above. Examples include recommendations on environmental health and on the health and care of the elderly. Categories were retained as standalone only when they accumulated more than 50 recommendations across the three UPR cycles; otherwise, they were grouped here.

**Decision rule:** Recommendations addressing violence against minorities, vulnerable groups, religious groups, human rights defenders, journalists, or asylum seekers that do not have an explicit health dimension are not considered health-related and are excluded from the dataset.

Categories considered but not applied

The category Social and economic determinants of health was considered during codebook development. Following team discussion, this category was not applied in the final classification, as recommendations addressing the social and economic determinants of health were judged to be most informatively classified under their specific thematic categories.

## Supplementary Figures

**Figure S1.**
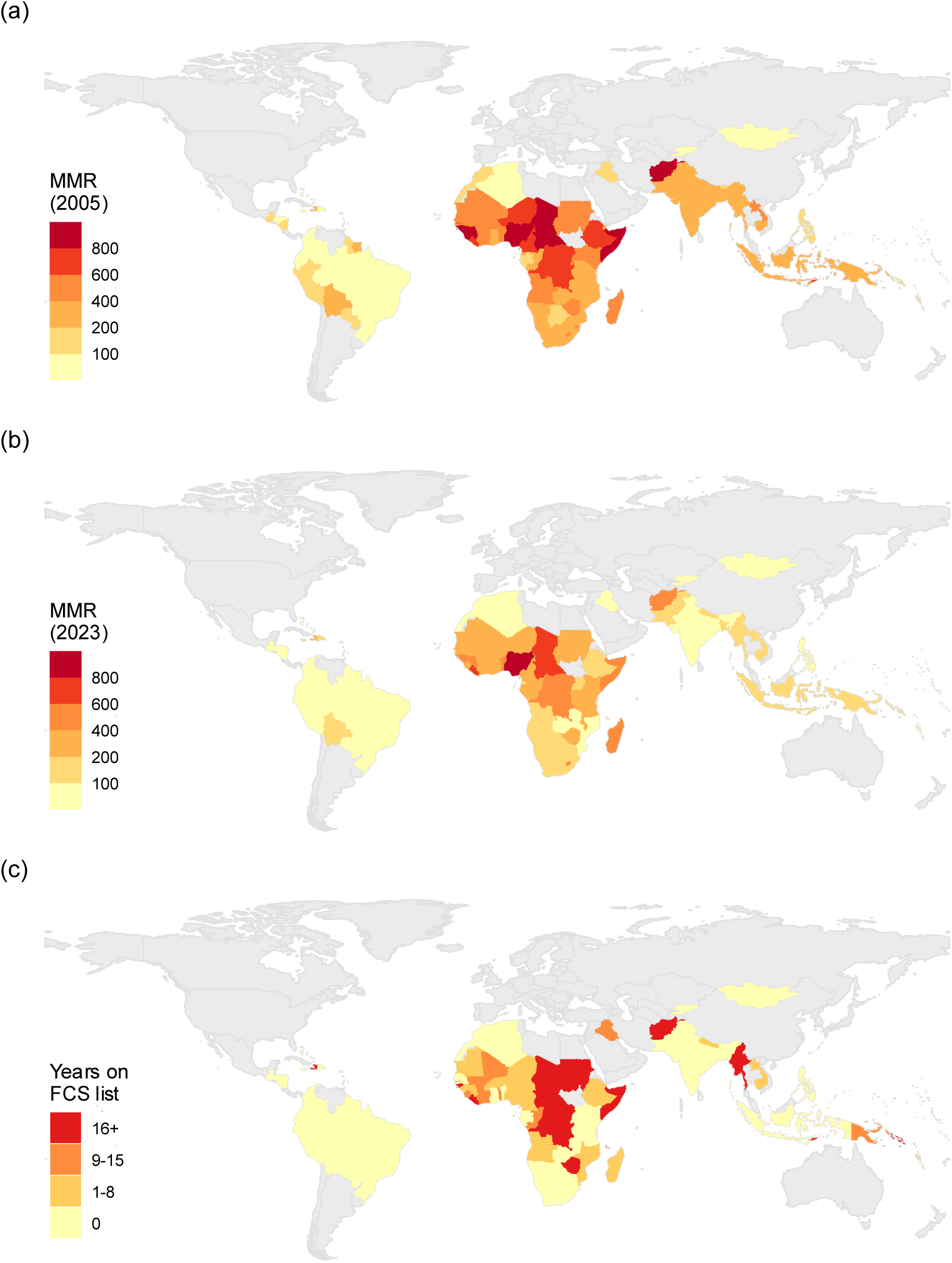
Map of analytical sample (n=89 countries) showing MMR estimates in (a) 2005 and (b) 2023, as well as (c) the cumulative number of years on the World Bank “Fragile and Conflict-affected Situations” (FCS) list, between 2006 and 2023.

**Figure S2.**
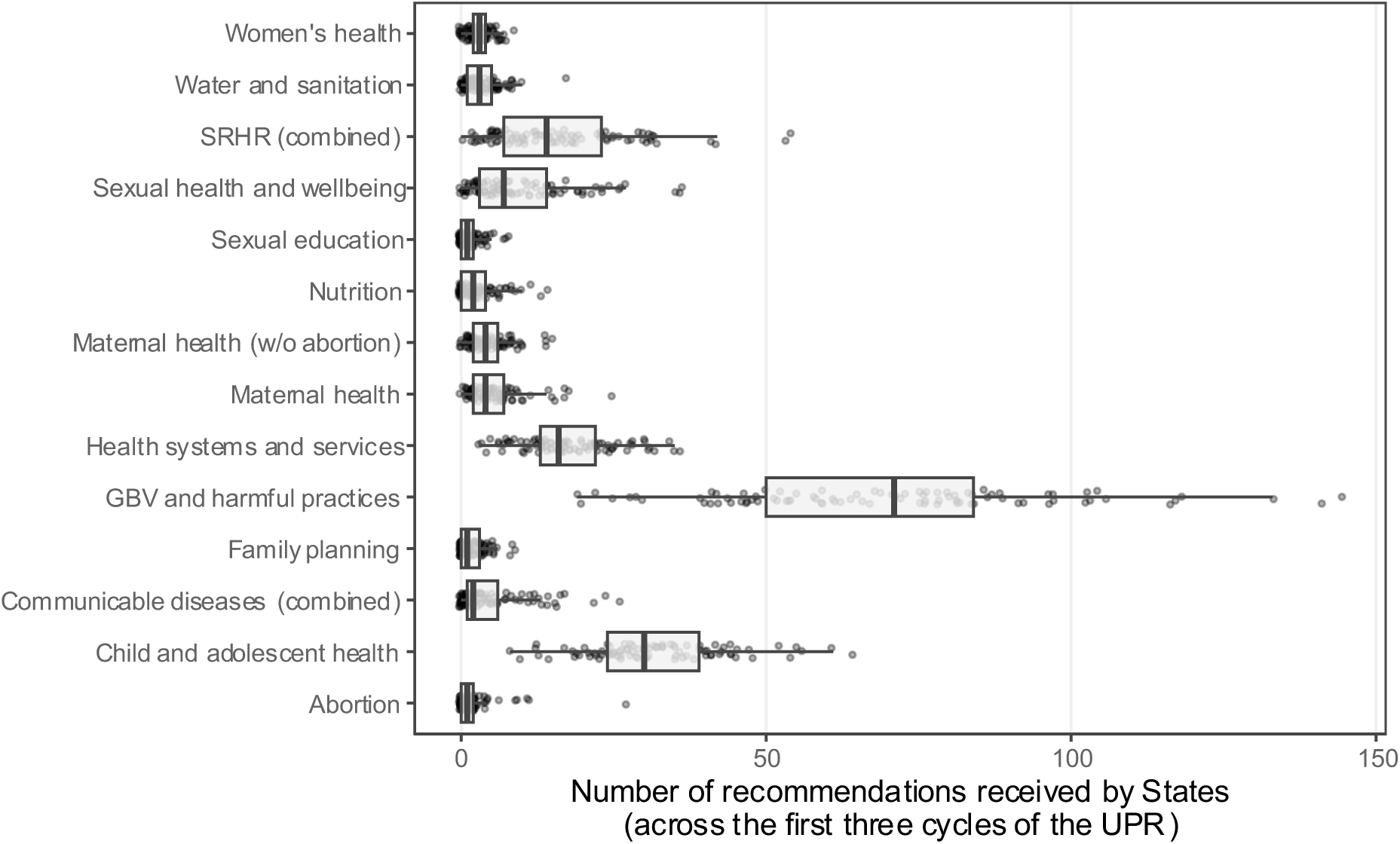
Boxplots showing the distribution of health-related UPR recommendations from the first three cycles of the UPR, by thematic grouping. Each point represents the number of recommendations that an individual country received.

## References

1. WHO. Trends in maternal mortality estimates 2000 to 2023: estimates by WHO, UNICEF, UNFPA, World Bank Group and UNDESA/Population Division. Geneva: World Health Organization; 2025.

2. Faith J, Martopullo I, Arndt MB, Letourneau ID, Aalruz H, Abaraogu UO, et al. Global, regional, and national levels and trends in maternal mortality, progress towards the Sustainable Development Goals, and mortality from COVID-19 infection in pregnant women, 1990–2023: a systematic analysis for the Global Burden of Disease Study 2023. The Lancet Obstetrics, Gynaecology, & Women’s Health. 2026; S3050503826000476. doi:10.1016/S3050-5038(26)00047-6

3. Alkema L, Chou D, Hogan D, Zhang S, Moller A-B, Gemmill A, et al. Global, regional, and national levels and trends in maternal mortality between 1990 and 2015, with scenario-based projections to 2030: a systematic analysis by the UN Maternal Mortality Estimation Inter-Agency Group. The Lancet. 2016;387: 462–474. doi:10.1016/S0140-6736(15)00838-7

4. WHO. Maternal mortality in fragile and conflict-affected situations: technical brief. Geneva; 2025. Available: https://www.who.int/publications/i/item/9789240115545/

5. Cresswell JA, Alexander M, Chong MYC, Link HM, Pejchinovska M, Gazeley U, et al. Global and regional causes of maternal deaths 2009–20: a WHO systematic analysis. The Lancet Global Health. 2025;13: e626–e634. doi:10.1016/S2214-109X(24)00560-6

6. Souza JP, Day LT, Rezende-Gomes AC, Zhang J, Mori R, Baguiya A, et al. A global analysis of the determinants of maternal health and transitions in maternal mortality. The Lancet Global Health. 2024;12: e306–e316. doi:10.1016/S2214-109X(23)00468-0

7. WHO. Strategies toward ending preventable maternal mortality (EPMM). 2015. Available: https://www.who.int/publications/i/item/9789241508483

8. Ronsmans C, Graham WJ. Maternal mortality: who, when, where, and why. The Lancet. 2006;368: 1189–1200. doi:10.1016/S0140-6736(06)69380-X

9. UN General Assembly. International Covenant on Economic, Social and Cultural Rights. Resolution 2200A (XXI) 1966.

10. Yamin AE. Applying human rights to maternal health: UN Technical Guidance on rights-based approaches. International Journal of Gynecology & Obstetrics. 2013;121: 190–193. doi:10.1016/j.ijgo.2013.01.002

11. WHO. Keeping promises, measuring results: Commission on information and accountability for Women’s and Children’s Health. World Health Organization; 2011.

12. OHCHR. Technical guidance on the application of a human rightsbased approach to the implementation of policies and programmes to reduce preventable maternal morbidity and mortality. Geneva; 2012. Report No.: A/HRC/21/22. Available: https://docs.un.org/A/HRC/21/22

13. Etone D, Nazir A, Storey A. Introduction. 1st ed. Human Rights and the UN Universal Periodic Review Mechanism. 1st ed. London: Routledge; 2024. pp. 1–9. doi:10.4324/9781003415992-1

14. Bueno De Mesquita J. The Universal Periodic Review: A Valuable New Procedure for the Right to Health? Health Hum Rights. 2019;21: 263–277.

15. Etone D. Theoretical challenges to understanding the potential impact of the Universal Periodic Review Mechanism: Revisiting theoretical approaches to state human rights compliance. Journal of Human Rights. 2019;18: 36–56. doi:10.1080/14754835.2019.1579639

16. Lane M. The Universal Periodic Review: A Catalyst for Domestic Mobilisation. Nordic Journal of Human Rights. 2022;40: 507–528. doi:10.1080/18918131.2022.2139076

17. Gilmore K, Mora L, Barragues A, Mikkelsen IK. The Universal Periodic Review: A Platform for Dialogue, Accountability, and Change on Sexual and Reproductive Health and Rights. Health Hum Rights. 2015;17: 167–179.

18. Yamin AE. Toward transformative accountability: applying rights-based approach to fulfill maternal health obligations. SUR-Int’l J on Hum Rts. 2010;7: 95.

19. WHO. Advancing the right to health through the universal periodic review. Geneva: World Health Organization; 2019. Available: https://iris.who.int/handle/10665/277114

20. UNFPA, Universal Rights Group. Advancing Rights, Transforming Lives: UNFPA strategic engagement with the United Nations human rights system to advance sexual and reproductive health and rights. New York: United Nations Population Fund; 2023. Available: https://www.universal-rights.org/urg-policy-reports/advancing-rights-transforming-lives-2/

21. Palmer A, Tomkinson J, Phung C, Ford N, Joffres M, Fernandes KA, et al. Does ratification of human-rights treaties have effects on population health? The Lancet. 2009;373: 1987–1992. doi:10.1016/S0140-6736(09)60231-2

22. Ahmed S, Ali M, Shah I, Tsui A. Effect of maternity care improvement, fertility decline, and contraceptive use on global maternal mortality reduction between 2000 and 2023: results from a decomposition analysis. The Lancet Global Health. 2025; S2214109X–25004097. doi:10.1016/S2214-109X(25)00409-7

23. WHO. Global health observatory data repository. 2026 [cited 2 Feb 2026]. Available: https://www.who.int/data/gho

24. UNFPA. Population Data Portal. In: Contraceptive prevalence rate, all women [Internet]. 2026 [cited 2 Feb 2026]. Available: https://pdp.unfpa.org/?page=Indicator-Overview#/viewer?indicator=588

25. OHCHR. Universal Human Rights Index. 2026 [cited 2 Feb 2026]. Available: https://uhri.ohchr.org/en

26. Charlesworth H, Larking E, editors. Human Rights and the Universal Periodic Review: Rituals and Ritualism. 1st ed. Cambridge University Press; 2015. doi:10.1017/CBO9781316091289

27. Gujadhur S, Limon M. Towards the third cycle of the UPR: stick or twist? : lessons learnt from the first ten years at Universal Periodic Review. Versoix: Universal Rights Group; 2016.

28. Meier BM, Onzivu W. The evolution of human rights in World Health Organization policy and the future of human rights through global health governance. Public Health. 2014;128: 179–187. doi:10.1016/j.puhe.2013.08.012

29. Meier BM, Bueno De Mesquita J, Sekalala S. Human Rights Limitations in World Health Organization Reforms: Strengthening Human Rights Obligations in Global Health Law to Ensure Global Health Equity. AJIL Unbound. 2026;120: 103–110. doi:10.1017/aju.2026.10058

30. Goodman R, Jinks D. How to influence states: Socialization and international human rights law. Duke lj. 2004;54: 621.

31. Barasa E, Chuma J, Nonvignon J, Adeyi OO. Avoidable pitfalls on the path to health financing self-reliance in low-income and middle-income countries. BMJ Glob Health. 2025;10: e021270. doi:10.1136/bmjgh-2025-021270

32. Wakefield J. Ecologic Studies Revisited. Annu Rev Public Health. 2008;29: 75–90. doi:10.1146/annurev.publhealth.29.020907.090821

33. Jamison DT, Summers LH, Alleyne G, Arrow KJ, Berkley S, Binagwaho A, et al. Global health 2035: a world converging within a generation. The Lancet. 2013;382: 1898–1955. doi:10.1016/S0140-6736(13)62105-4

